# Application and content of minimum data sets for care homes: A mapping review

**DOI:** 10.1101/2024.06.24.24309361

**Authors:** Barbara Hanratty, Gizdem Akdur, Jennifer Kirsty Burton, Vanessa Davey, Claire Goodman, Adam Lee Gordon, Anne Killett, Jenny Liddle, Stacey Rand, Karen Spilsbury, Ann-Marie Towers

## Abstract

**Background:** Care home residents have complex needs, and minimum data sets (MDSs) provide a unique source of information on their health and wellbeing. Although MDSs were first developed to monitor quality and costs of care, they can make an important contribution to research.

**Aim:** To describe the research applications of data from care home MDSs, and identify key outcome variables and measures used.

**Design:** Mapping review of published empirical studies using data generated from minimum data sets in long term care facilities for older adults.

**Methods:** We performed a comprehensive search of electronic databases (Medline OVID, CINAHL, Embase and ASSIA), using bespoke search strategies to identify English language publications 2011 - 2024. Articles were screened by two independent reviewers. They were grouped by study topic and data (on publication date, country, MDS, outcome variables and specific items or measures) were charted without quality assessment. The key features of the data are described in a narrative synthesis.

**Findings:** Searches identified 18588 articles published 2011-2024, of which 661 met inclusion criteria. 72% were from the USA, 12% from Canada and the remaining 16% from four European countries, South Korea and New Zealand. The studies encompassed individual resident functioning (e.g. mobility, incontinence), health conditions and symptoms (e.g. depression, pain), healthcare in the home (e.g. prescribing, end of life care), hospital attendances and admissions, transitions to and from care homes, quality of care and systemwide issues. Measures used reflected the content of the major MDSs, but there was a mismatch between the importance of some topics to care homes (e.g. incontinence) and the range of published papers, and limited consensus over how to measure quality of life.

**Conclusions:** Care home MDSs are a unique resource to support study of care home residents and impact of interventions over time. They are a powerful resource when linked to other datasets, and as an adjunct to primary data collection This analysis may serve as an accessible guide to the content and applications of MDS, allowing researchers to consider the sort of questions that can be posed and the different components of resident care or experience that can be evaluated.

## Introduction

Care home residents have complex needs, and a good understanding of their health and wellbeing is the key to optimising care.^1^ ^2^ This is an important group, growing in size and with increasing needs for care.^3^ ^4^ Over the next twenty years, the number of people requiring the level of support provided by a care home is expected to increase by 50%, in line with population ageing.^5^ ^6^ Research from Europe, New Zealand and North America has described rising care needs and acuity of medical conditions amongst care home entrants.^7^ ^8^ Older age, cognitive impairment and difficulties with activities of daily living (ADLs) are established risk factors for care home admission.^9^ Three out of four residents have reduced mobility and a similar proportion live with incontinence.^10^ Arthritis, depression and dementia are common,^1^ ^10^ and an increasing number of residents are admitted directly from hospitals, with associated needs for rehabilitation and infection control.^6^ Demands on health services may be high, with greater use of primary care by older adults in care homes, compared to people in their own homes, more frequent hospital attendances^10^ and high rates of in-hospital death.^11^

A minimum data set (MDS) describes a core dataset, collected for a specific purpose. Most often, MDSs are focused on demographic, social, economic or health characteristics. Care home MDSs may provide data on individual residents, staff and homes, which can be aggregated to study activity at care home, organisation or population levels. Over time, the resulting longitudinal data on residents’ and their home can be a resource to support planning, delivery and evaluation of services tailored to residents’ needs.^12^ An MDS can also enable national reporting on care metrics, cross-national comparative studies, and - in conjunction with other healthcare record systems - studies of how older adults move between care homes, hospitals and the wider community.^13^ ^14^ An MDS for care homes has been used in the USA for almost 30 years, where Medicaid and Medicare affiliated care homes are now mandated to use the it (currently version 3.0).^15^ Two other forms of MDS, the Resident Assessment Instrument and Dutch National Measurement of Care Problems (LPZ) ^16–20^ are in use elsewhere. All MDSs specify the schedule and content of assessments, support standardised data collection, and provide protocols to guide assessment, and in some cases, triggers for escalation of care. Established MDS are lengthy tools, with the US MDS version 3.0 consisting of over 400 items.^15^ They form part of a vast array of tools, available to use with care home populations. A recent review of quality management guidelines and frameworks for care homes from seven European countries and the US identified 94 performance indicators across five domains. The quality of care domain alone included 24 indicators.^21^

Many countries have no regular sources of data from care homes and no reliable means of identifying care home residents within routine data sources.^22^ The coronavirus pandemic that started in 2020 highlighted the importance of this omission, and the potential benefits of data collection for residents, care systems and research. Data on activity and COVID-19 infection in care homes were needed urgently, but infrastructure to support data collection and transfer were often absent. In countries without MDS or similar systems, there are examples of swift action to redress this information gap. For example, in England, a capacity tracker app was used to provide aggregate information such as care home bed occupancy and staffing, to local authorities.^23^ This and other interventions during the pandemic, heightened interest in the potential of MDS to support responses to emergencies and rational planning, as well as research.

The purpose of this study was twofold. First, to provide an overview of the breadth of topics that may be studied using data from care home MDS. Second, to map the key measures used within each topic area. We did this to generate a resource for researchers, with a guide to where MDS has supported research, and where the gaps are.

## Methods

We undertook a mapping review of the published literature. This was judged to be the most appropriate approach, as we were interested in a broad overview of the content of a large literature. It allows researchers to map and describe the literature in a specific area, and highlight important concepts.^24^ We adopted the established standards for the conduct of these type of reviews. ^25,26–28^ In keeping with the mapping review approach, we extracted limited, descriptive information about studies. The purpose was to understand how MDS data has been used in research and identify items/measures used in common topic areas of MDS research. Throughout this review, we use the terms ‘items/variables’ to describe responses to single items or questions and ‘measures’ to describe multiple items (more than two) forming a measurement scale.

### Search Strategy

This work utilised broad searches developed for a linked review.^29^ Initially, a bespoke search strategy was developed with an information scientist to identify original studies analysing data from care home MDSs. It was used to search seven data bases (Medline OVID, SCOPUS, CINAHL, Embase, ASSIA) from 2011 to 2015, subsequently updated to 2019.^29^ (Search strategies reported elsewhere and reproduced in the Supplementary Material).^29^ Further updating was undertaken to 2024. This provides fourteen full years of publications, with recent data most likely to reflect current practice. Additional targeted searches used the names of MDSs known to be implemented (e.g. MDS, InterRAI, LPZ) and by manually searching the reference lists of potentially relevant papers and sources of grey literature, including websites of MDS providers. References were imported into Endnote software for screening.

### Inclusion and exclusion criteria

Studies were included if they reported original research, using data from an MDS in care homes (care home, residential home, long-term care facility, skilled nursing facility). Studies that described development of an MDS, validated measures within MDSs, or used measures taken from MDSs to undertake data collection, were all excluded. We included articles that combined analysis of previously collected MDS data with new data collection in a single study. No restrictions on language, country, age of study participants, type of care home or study topic were applied at this stage. All observational and experimental study designs were eligible, but editorials, reviews, commentaries and conference abstracts were excluded. Included studies were published between January 2011 and March 2024, to ensure the findings reflected recent practice in care homes.

### Title and abstract, full text screening

Title, abstract and full text screening were all completed in EndNote. One reviewer screened all records, with 20% checked by a second reviewer. All uncertainties were resolved by discussion between two reviewers, with a third reviewer available to intervene in the event of conflicts. Studies that presented analysis of MDS data were included for data extraction.

### Data charting

A data charting form was developed in Microsoft Excel, tested on ten articles and modified before applying to all included studies. Descriptive data were charted from each article that met the inclusion criteria: year of publication, country, study topic, MDS, and items or measures in the topic area being studied.

Given the study heterogeneity and breadth of topics, a narrative synthesis approach was used to collate and summarise the literature, including a count of study characteristics. Articles were categorised by study topic, with some studies covering more than one category. The study categories were then grouped into broader thematic groups, based on our interpretation of the categories. No assessment of study quality was made, as this was a mapping review.

### Findings

After deduplication, searches identified 18,588 articles published 2011-2024, of which 661 met inclusion criteria. The majority (72%) were from the USA, 12% Canada and the remaining 16% from four European countries, South Korea and New Zealand. The average annual number of relevant publications was 51 with a peak in 2020 (90 articles). As the overall number of studies in the review is high, the narrative below includes only selected references. Complete lists, by category, are provided in the supplementary material. An adapted PRISMA diagram is presented in Figure 1.^30^

**Figure 1.** (PRISMA diagram)

#### Topics studied using information from MDS

Overall, one of the most common uses of MDSs in this review was to describe residents, either in epidemiological terms, as described below, or in terms of their health, functioning and service use. This review includes studies that have considered individual resident functioning (e.g. mobility, incontinence), health conditions and symptoms (e.g. depression, pain), health care in the home (e.g. prescribing, end of life care), hospital attendances and admissions, transitions to and from care homes, quality of care and system-wide issues. Information from MDSs has been particularly useful when employed in empirical studies, providing a sampling frame and baseline for randomised controlled trials or other intervention studies.^31–33^ Many studies have used MDS data linked to other sources of information, most commonly routine data sets such as those collected by Medicare. This provides a comprehensive picture of the resident’s care as they transit across settings. It also enables assessment of outcomes that may be important to residents in their daily lives (e.g. activities of daily living), but may not be the intended primary outcome for medical care.

**Table 1.** Study topics in research using MDS*.

| TOPIC | SUB-CATEGORIES | NUMBER OF ARTICLES |
| --- | --- | --- |
| **Epidemiology & research** | Describing residents | 87 |
|  | MDS as a supplement to primary study | 61 |
|  | Research methods | 23 |
|  | Mortality risk prediction | 15 |
|  | Prediction of other outcomes | 32 |
|  | Preventive approaches | 75 |
|  | Outcomes | 34 |
| **System-wide issues** | Care home admissions | 20 |
|  | Care home bed use | 17 |
|  | Hospital admissions & readmissions | 40 |
|  | Emergency department attendance | 16 |
|  | Post-acute care | 37 |
|  | Staffing | 4 |
|  | Trends | 12 |
| **Quality** | Quality of care | 71 |
|  | Quality of life | 13 |
|  | Resident preferences | 5 |
| **Disability and impairment** | Functional assessment | 53 |
|  | Incontinence | 19 |
|  | Falls | 32 |
|  | Fracture prevention | 22 |
| **Health** | Mental health | 16 |
|  | Dementia | 87 |
|  | Cognitive impairment | 26 |
|  | Depression | 9 |
|  | Delirium | 7 |
|  | Frailty | 32 |
|  | Obesity and nutrition | 10 |
|  | Multimorbidity | 5 |
|  | Parkinson's | 3 |
|  | Infections | 39 |
| **Symptoms** | Pain | 39 |
| **Prescribed medications** | Medication | 121 |
|  | Antidepressants | 12 |
|  | Anticoagulants | 7 |
|  | Anticholinergics | 8 |
|  | Antibiotics | 12 |
|  | Antipsychotics | 40 |
|  | Anxiolytics | 4 |
|  | Diabetes medications | 17 |
|  | Deprescribing | 12 |
| **End of life care** | End of life care | 51 |
|  | Advanced care planning | 2 |

- Numbers will not sum to total included articles, as many fall into more than one group,

##### Health conditions

MDSs can support investigation of the epidemiology, care and outcomes for specific health conditions amongst residents, despite not being healthcare records. Dementia and cognitive impairment are amongst the most commonly studied conditions in this regard. Mental health has also been an important area of study,^34^ particularly the treatment of depression^35^ and anxiety.^36^ Prediction, prevention and treatment of infections are also feasible applications of MDSs. For example, studies have looked at trends in infections over time,^37^ risk factors for developing pneumonia,^38^ evaluation^39^ and use^40^ of vaccines. MDSs have been used to investigate specific infections that have serious consequences for older adult residents, such as clostridium difficile,^41^ norovirus^42^ or methicillin resistant staphylococcus aureus.^43^

##### Medications

Analysis of medications prescribed in care homes was the most common application of data from MDS in this review. De-prescribing or de-intensification of treatment was the explicit focus of a modest number of studies.^44–53^ Research also examined specific categories of medications with known potential adverse effects for older adults, such as those with a high anticholinergic burden,^54–61^ anticoagulants,^62–68^ antidepressants,^69–80^ anti-psychotics,^31^ ^36^ ^44^ ^70^ ^77^ ^78^ ^81–107^ or statins.^49–51^ ^108–112^ Analysis of prescribing in some medication categories - antibiotics^113–116^ or benzodiazepines,^84^ ^117^ for example, may be used to make broader assumptions about quality of care. A common approach in MDS-based research has been to consider medications as risk factors for adverse events in care homes. These have included, for example, falls,^47^ ^70^ ^96^ ^118^ fractures,^73^ ^101^ ^119–121^ frailty^122^ and death.^57^ ^69 90 92 94 104 123^

##### Pain

Assessment and prevalence of pain have been extensively studied using MDSs,^124^ ^125^ along with pharmacological management.^126^ ^127^ Authors have also explored factors associated with pain, including cognitive impairment^128^and dementia, and the consequences for behaviour within the home.^129–131^

##### Assessment of functional status or decline

The ability of an older adult to perform day to day tasks, such as washing and dressing, and their mobility, have important implications in care homes. ADLs are available from MDS, but seldom recorded in healthcare records. Researchers have used MDS to explore changes in functional ability over time,^132–135^ after acute medical and surgical hospital admissions,^136^ ^137^ ^138^ ^139^ and for residents who are ultimately discharged home with^140^ or without restorative (rehabilitative) care.^137^

##### Falls

Falls are recorded in care homes across the world as a measure of care quality and safety, and often as a regulatory requirement. Falls data from MDS have been used to characterise the scale and consequences,^141–143^ and identify potential risk factors for falls, including functional ability,^118^ health status^144^ ^145^ and a range of medications.^47^ ^70^ ^96^ ^146^ Information on falls has also been analysed as part of wider research studies^147^ ^148^ and supplemented with data from healthcare records.^149^

##### COVID-19

The coronavirus pandemic had broad ranging consequences for staff and resident health and wellbeing, as well as the organisation, delivery, regulation and scrutiny of care. This is reflected in the analyses of MDS, though the number of studies with a particular focus on COVID-19 was modest.^150–156^

##### Hospital admissions

Across the world health systems strive to ensure that admissions to hospital from care homes only occur if appropriate and unavoidable. MDSs have been a common source of information for research on admissions^157^ ^158^ and readmissions. Individual and institutional factors associated with admissions have been studied, including mental and physical health,^159^ ^160^functioning,^161^ race,^162^ payer status, quality of care in the home,^163^ and ownership status of the care home.^164^

##### Neglected topics

There are a number of topics, such as incontinence, oral health, that are important to residents and staff in care homes, but often neglected by research. We found a small number of studies concerned with the management or sequelae of incontinence,^165–183^ but few with a specific focus on faecal incontinence.^170^ ^174^ ^177^ In this review, a majority of the identified papers on dental or oral health were excluded because they focused on validating measures, rather than analysing data collected within an MDS. Those that were included all used the MDS RAI oral/dental items.^184^ ^32^

#### Measures and items

Almost all studies in this review utilised MDS 3.0 (or preceding versions) or RAI and its versions. Hence, the majority of measures in this review were drawn from these well-established tools.

#### Quality of Life

There is widespread acknowledgement of the importance of resident-level quality of life (QoL) in care homes. QoL as it reflects the quality, safety and effectiveness of care from the resident’s perspective (‘care-related quality of life’) is important, but also residents’ health-related and condition-specific QoL. However, the importance of the topic does not appear to be reflected in the breadth of published papers using routinely collected data to investigate QoL. This may be partly due to the availability of QoL data from established MDS. InterRai’s quality of life survey contains 49 items, but they are self-reported, and they may be liable to poor completion. Other domains of QoL that are important to residents and reflect the quality of care, e.g. interpersonal relationships may be difficult to capture.^185^ Some of the dimensions that have been labelled as QoL could also be appropriately considered to be basic needs – e.g. safety and security, food, comfort, access to services. The lack of clarity over the measurement of QoL is also exemplified in another study, which measured changes in functional status, weight and depressive symptoms, and termed them QoL outcomes.^152^ Measures of resident-level QoL (e.g. the Minnesota Quality of Life Survey) have been implemented in care homes in some geographical locations, but this has been to supplement collection of information for MDS, and compare to different MDS components.^186^ ^187^ Such localised data collections have enabled studies that examine the association between individual and institutional factors and QoL over time.^188^

## Discussion

This review has demonstrated the potential of MDSs to contribute to enhancing care and outcomes for care home residents through research. The research application of data from care home MDSs is broad. Key categories studied were description of resident status (e.g. functional assessment), symptoms (e.g. pain), service use (e.g. general and care of specific diagnoses or points in the life course), prescribing and quality of care. A wide range of measures were used, but the majority reflected the content of contemporary, implemented MDS.

The value and application of MDSs is increased by linking to other sources of routine information. In this review, many of the studies analysing MDSs linked to health service datasets. Such enlarged data sources support scrutiny of transitions across settings, evaluation of outcomes of care or generation of an in-depth picture of care delivery. Data linkage may also be helpful to identify care home residents in other routine data sources. This is an ongoing challenge in many countries, that inhibits basic study of care home resident health and social care utilisation and patterns of mortality.^189^ ^190^

There are notable gaps in the published MDS research on issues that are important to residents and families, such as QoL, incontinence and oral health. There is an emerging focus and consensus on measurement of QoL in this setting, which could inform the future addition of QoL measures, especially for research purposes. Common metrics across datasets and studies would support comparative research and may act as a stimulus for intervention development. Previous studies have taken a broad interpretation of the term ‘QoL’, which may be useful, but often leads to imprecision in what is being measured and a diversity of QoL measures used. Future MDS research ought to carefully consider the construct of interest (e.g. care-related, health-related or condition-specific QoL), as well as the study’s specific aims and objectives and the emerging consensus (add DACHA refs), to guide the selection of QoL measure(s) for their analysis.

### Comparison with other work

A previous review of measurement tools designed for the management and care of older people in long term care identified 25 instruments. Similar to our study, a majority were drawn from MDS and RAI-MDS. Depression, cognition and functional capacity were the most commonly assessed domains.^191^ Our review was broader because it included measures used in published research studies, some of which were developed in different settings. A common finding from both reviews is the clear potential of MDS to make a significant contribution beyond research, to the practical aspects of care home life.^192^ The role of MDS in quality improvement activities has been well documented. Previous work showed that the Resident Assessment Instrument has been employed most often in the development of process improvement models, and in multi-faceted approaches and educational interventions to improve the quality of care.^193^ The absence of any measurable impact was attributed to a paucity of staff and time, and turnover of personnel. End of life care is one of the most important aspects of care home work, but one that presents particular challenges to the measurement of quality. Eleven measures used in long term care settings to assess the quality of care when a resident is dying were identified by van Soest-Poortvliet and colleagues.^194^ Key areas covered by the 11 instruments (340 items) were structure, process and evaluation of care (42%) and the quality of dying (38%). Most measures (7/11) encompassed multiple constructs, but it is noteworthy that the two developed outside of North America placed far less emphasis on measuring processes of care. A recent review of outcome measures from intervention research with care homes found similar gaps in under-studied areas, such as incontinence, but more studies on health-related QoL.^195^ This latter may be explained by our focus on studies that analysed routinely collected data, rather than those that validated or tested QoL measures.

Our review focused on the content of MDS, and how the data may be applied to resident care and outcomes. Analyses of large datasets may be criticized as insensitive to resident experiences and perceptions. The US MDS 3.0 strives to overcome this by requiring care homes to interview residents with scripted questions to assess pain, mood and cognitive functioning. In practice, although a high proportion of long stay residents do participate, people in smaller homes and those with cognitive impairment or in the last six months of life are less likely to have their voices heard.^196^

### Strengths and limitations

This is a novel review of the content and use of care home MDS data. Mapping of items and measures used in research on different topics can inform the development of future MDS. The review has also generated a resource for researchers, by highlighting the possible uses of MDS data, and areas where research is limited, including due to absence of data collection (e.g. QoL). However, it should be acknowledged that a review of this size is prone to errors and omissions. We excluded studies that used MDS derived items or measures to collect data, or implemented MDS in a research context. This gave prominence to studies from the USA and Canada where MDS are well established. Financial incentives and federally mandated data collection in the USA are a key concern. In other countries, statutes and regulations have a role in implementing standardised assessments,^197^ and it is important to consider the local context when drawing conclusions from cross national studies. A number of European studies have made a major contribution to our understanding of the feasibility and potential of MDS. Notably, the Services and Health for Elderly in Long TERm care study (SHELTER) assessed the validity of InterRAI in seven European Union countries and Israel.^198^ This research generated findings across a range of topic areas including prescribing, sensory impairment, prognostication and sleep. ^100^ ^198–216^ ^205^ ^217^ ^218^ In Italy the ULISSE study involved 31 nursing homes in a study of the quality of care delivered to older people, using InterRAI based assessments. ^219–222^ Whilst excluding these and other research initiatives from our review may limit the generalisability of our findings, it allows us to answer questions about what can be achieved with MDS data collected in a real-world context, without research staff input.

### Implications and conclusions

MDS are an invaluable source of longitudinal data on care home residents, generating unique insights into health, wellbeing and functional status. For researchers there are two scenarios where MDS are particularly useful. First, when linked to other datasets, MDSs provide a powerful resource for investigation of factors that may be associated with positive and adverse outcomes, including transfer to hospital and mortality. The range of factors that can be investigated is vast, ranging from funding models to institutional and individual characteristics. However, availability of such rich data within the MDS makes it critically important that researchers ensure that they have a robust hypothesis or rationale for their analyses. Second, MDSs can contribute to empirical studies involving primary data collection. Generating baseline data from an MDS, or providing a comparison group for an intervention study helps to keep research costs down by reducing the work of data collection. Modern analytical techniques combined with MDSs offer exciting possibilities to

There are some important gaps in the body of MDS research that should be addressed. QoL has often been conceptualised in terms of health and functioning, rather than potentially more holistic, social care related outcomes. In addition, where MDS use is established, there may be limited ability to modify or influence the data collected. Other apparent gaps – such as decline in functional status, sensory impairment and incontinence – point to a need for MDS based researchers to ensure their questions reflect the priorities of residents and those who care for them. This is important, because when MDS data does reflect the experience of care, data entry is improved, and the information can be used as a basis for care planning and review.^223^

## Data Availability

All data produced are contained in the manuscript. This is an analysis of material already in the public domain.

