## Supplementary material for "Application and content of minimum data sets for care homes: A mapping review"

**Figure 1: Prisma Diagram**

**Identification of studies via databases and registers**

Records removed *before screening*:

Duplicate records removed (n = 1245)

Records identified from databases (n=19,833)

**Identification**

Records screened

(n = 18588)

Records excluded**

(n = 17851)

Reports sought for retrieval

(n = 773)

Reports not retrieved

(n = 0)

**Screening**

Reports excluded:

Not using MDS data (n = 14)

Wrong population or setting (n = 3)

Primary studies collecting data (n = 28)

Review article (n =6 )

Validation study for measure or tool (n = 13)

Commentary or other design with no data (n =18 )

SHELTER & ULISSE studies (n=19)

Other reasons (n= 11)

14

Reports assessed for eligibility

(n = 773)

Studies included in review

(n =661)

**Included**

**Search Strategies**

**Medline search strategy (adapted for other databases, and supplemented with MDS specific searches)**

1. *Homes for the Aged/

2. *Nursing Homes/

3. *Long-Term Care/

4. *Residential Facilities/

5. *Respite Care/

6. *Intermediate Care/

7. "care home$".ab,ti.

8. "nursing home$".ab,ti.

9. "residential care".ab,ti.

10. ("long term care" or "long-term care" or "longterm care").ab,ti.

11. "home$ for the aged".ab,ti.

12. "care facilit*".ab,ti.

13. "old$ people$ home$".ti,ab.

14. (retir$ adj2 home$).ab,ti.

15. ("old$ adult$" adj3 (facilit$ or residential or accommodation)).ab,ti.

16. ("old$ people$" adj3 (facilit$ or residential or accommodation)).ab,ti.

17. ("old$ person$" adj3 (facilit$ or residential or accommodation)).ab,ti.

18. ((geriatric$ or elder$ or senior$ or retir$) adj3 (facilit$ or residential or accommodation)).ab,ti.

19. "respite care".ti,ab.

20. "intermediate care".ti,ab.

21. or/1-20

22. *Randomized Controlled Trials as Topic/

23. Randomized controlled trial/

24. Random allocation/

25. Double blind method/

26. Single blind method/

27. Clinical Trial/

28. Clinical trials as Topic/

29. "randomi*ed".ab,ti.

30. randomly.ab,ti.

31. controlled clinical trial.pt.

32. Evaluation Study/

33. Comparative Study/

34. "before and after study".ti,ab,mp.

35. or/22-34

36. 21 and 35

37. limit 36 to yr="2009 -Current"
